# Metformin Use, Inflammatory Markers, and Mortality in U.S. Adults with Type 2 Diabetes: A Nationally Representative Cohort Study Using NHANES 2013–2018

**DOI:** 10.64898/2026.04.29.26352069

**Authors:** Yizhu Chen, Jinghao Guan, Yidong Wang, Yifan Xu, Haoxiong Sun

**Affiliations:** School of Pharmacy, University College London, London, United Kingdom; National Heart and Lung Institute, Imperial College London, London, United Kingdom; Division of surgery and interventional science, faculty of medical science, University College London; University of Manchester; Department of Behavioural Science and Health, University College London

## Abstract

Metformin has been linked to mortality benefits in type 2 diabetes that may extend beyond glycemic control, but population-level evidence connecting these benefits to inflammation-related pathways remains limited. Using NHANES 2013–2018 data with mortality follow-up through 2019, we examined associations between metformin use and four inflammatory markers, including neutrophil-to-lymphocyte ratio (NLR), monocyte-to-lymphocyte ratio (MLR), serum albumin, and high-sensitivity C-reactive protein (hs-CRP), and evaluated their relevance to all-cause and cardiovascular mortality. Among 2,122 adults with self-reported diabetes (60% metformin users; 2,116 with valid mortality follow-up), survey-weighted linear regression adjusted for demographic, socioeconomic, and metabolic covariates showed metformin use was associated with lower NLR (β = - 0.35; 95% CI −0.57, −0.14), lower MLR (β = −0.04; 95% CI −0.06, −0.02), and higher serum albumin (β = +0.11 g/dL; 95% CI 0.06, 0.16); the hs-CRP association was directionally consistent but not significant. Associations for NLR and MLR were essentially unchanged after BMI and HbA1c adjustment, remained robust in an active comparator analysis against sulfonylurea monotherapy, and were consistent across propensity score and overlap weighting sensitivity analyses. Survey-weighted Cox regression linked metformin to lower all-cause (HR 0.64; 95% CI 0.48, 0.86) and cardiovascular mortality (HR 0.49; 95% CI 0.26, 0.94). NLR was independently associated with all-cause mortality, with the highest tertile carrying nearly twice the hazard of the lowest, and inclusion of NLR or MLR modestly attenuated the metformin–mortality association. Metformin use is associated with a distinct cellular immune-inflammation profile in adults with type 2 diabetes, supporting further investigation of non-glycemic pathways relevant to its long-recognized clinical benefits.

## Introduction

Metformin is the most widely prescribed first-line therapy for type 2 diabetes mellitus. Landmark trial evidence from the UK Prospective Diabetes Study suggested that its clinical benefits extend beyond improvements in HbA1c, particularly in reducing macrovascular outcomes and all-cause mortality [1, 2]. Experimental studies have since proposed several non-glycemic mechanisms, including AMPK-dependent suppression of NF-κB signalling and modulation of innate immune function, which together implicate metformin in the attenuation of chronic low-grade inflammation [3, 4]. However, population-level evidence supporting these inflammation-related pathways remains limited, and it is unclear whether they translate into measurable differences in routinely available clinical markers among people with diabetes.

Complete blood count (CBC)-derived indices, including the neutrophil-to-lymphocyte ratio (NLR) and monocyte-to-lymphocyte ratio (MLR), are increasingly recognised as accessible markers of systemic immune-inflammation status. NLR reflects the balance between innate and adaptive immunity and has been shown to predict cardiovascular events and all-cause mortality in diabetic populations, while MLR captures monocyte-driven inflammatory activity relevant to atherosclerosis and vascular complications, which has been linked to diabetic complications [5–7]. Serum albumin, a negative acute-phase protein, and high-sensitivity C-reactive protein (hs-CRP), a positive acute-phase protein, serve as complementary indicators of chronic inflammatory burden that are routinely measured in clinical practice [8, 9]. Importantly, these markers reflect **partially distinct inflammatory pathways**: CBC-derived indices capture cellular immune composition, whereas albumin and hs-CRP reflect hepatic acute-phase responses closely linked to adiposity and metabolic status [10]. This distinction is methodologically relevant, as it allows inflammation-related drug effects to be probed across complementary biological axes using data already available in standard clinical panels.

Several studies have addressed aspects of the relationship between metformin use, inflammation, and mortality, but important gaps remain. A clinical cohort study in Scotland demonstrated that metformin users had lower NLR compared with sulfonylurea users [11], providing direct evidence linking metformin to cellular immune-inflammation differences. However, this finding was derived from a single-centre clinical population and lacked mortality follow-up, leaving it unclear whether the observed NLR difference carries prognostic significance. Separately, a population-based analysis of U.S. adults with diabetes established NLR as an independent predictor of all-cause mortality [6], but did not examine medication-specific associations, so the connection between metformin use and NLR-related mortality risk was not explored. More recently, serum albumin was identified as a factor partially accounting for metformin-associated mortality differences in NHANES [12], but this analysis was restricted to elderly patients with hypertension and did not incorporate CBC-derived immune indices such as NLR or MLR. Taken together, these studies each address one piece of the puzzle - the metformin–NLR association, NLR–mortality prediction, or albumin-related mortality mechanisms - but no study has simultaneously examined metformin’s association with CBC-derived immune-inflammation indices and assessed whether these indices contribute to explaining metformin-associated mortality differences in a nationally representative diabetic cohort.

The National Health and Nutrition Examination Survey (NHANES) is a nationally representative, continuously conducted survey of the U.S. population that collects detailed information on prescription medication use alongside a standardised set of clinical biomarkers, and can be linked to the National Death Index for longitudinal mortality follow-up. Using NHANES 2013–2018 data linked to mortality records through 2019, this study aimed to: (1) examine associations between metformin use and four complementary inflammation-related markers (NLR, MLR, serum albumin, and hs-CRP) in U.S. adults with diabetes, using both the general non-user comparison and an active comparator analysis against sulfonylurea monotherapy to mitigate confounding by indication; (2) evaluate whether these associations are accounted for by metabolic factors (BMI and HbA1c), thereby distinguishing inflammation-related markers whose differences with metformin use operate through metabolic versus non-metabolic pathways; and (3) assess the relevance of these markers to metformin-associated mortality differences, by examining whether their inclusion attenuates the metformin–mortality association through sequential adjustment, evaluating NLR as an independent mortality predictor in this population, and exploring potential effect modification of the metformin–mortality association across NLR strata.

## Methods

### Study Design and Data Source

We used data from three consecutive cycles of the National Health and Nutrition Examination Survey (NHANES 2013–2018), a nationally representative survey of the civilian, non-institutionalized U.S. population conducted by the National Center for Health Statistics using a stratified, multistage probability sampling design [13]. Cross-sectional analyses of inflammatory markers were combined with prospective mortality follow-up through the NHANES Public-Use Linked Mortality Files (through December 31, 2019) [14].

Adults aged ≥20 years with self-reported diagnosed diabetes were eligible. We excluded participants with hs-CRP greater than 10 mg/L, a threshold commonly used to minimize the influence of acute infection, as well as those with missing or zero MEC examination sample weights [15]. Details of inclusion and exclusion were shown in **Supplementary figure 1.** The cross-sectional analytic sample comprised 2,122 participants; of these, 2,116 had valid mortality follow-up time and constituted the prospective mortality cohort.

### Exposure and Outcomes

Metformin use was identified from the prescription medication questionnaire and was defined as any metformin-containing prescription reported within the previous 30 days. Concomitant use of sulfonylureas, insulin, and DPP-4 inhibitors was identified using the same approach for use as covariates and in sensitivity analyses.

Four inflammatory markers were examined: serum albumin, NLR, MLR, and hs-CRP. NLR was calculated as the absolute neutrophil count divided by the absolute lymphocyte count, and MLR was the absolute monocyte count divided by the absolute lymphocyte count. They were both derived from complete blood count differentials. Because hs-CRP was unavailable for the 2013–2014 cycle owing to a laboratory protocol change (approximately 37% missing), it was treated as a secondary outcome.

All-cause and cardiovascular mortality were determined through probabilistic linkage to the National Death Index. Cardiovascular mortality was defined as deaths from heart disease or cerebrovascular disease according to the NCHS underlying-cause groups. Survival time was measured from the MEC examination date.

### Covariates

Covariates were selected a priori based on established associations with both metformin use and inflammation. They were grouped as demographic (age, sex, race/ethnicity), socioeconomic and behavioral (education, poverty-income ratio, smoking), and metabolic (BMI, HbA1c). Enhanced covariates used in sensitivity analyses additionally included physical activity and alcohol use.

### Statistical Analysis

All analyses incorporated the NHANES complex survey design, including primary sampling units, stratification, and MEC examination sample weights. The original two-year cycle weights were divided by 3 to construct 6-year pooled examination weights for the combined 2013–2018 sample. All tests were two-sided at α = 0.05, and analyses were performed in R version 4.5.0.

Cross-sectional associations were estimated using survey-weighted linear regression with three adjusted models: demographic factors only (Model 1); Model 1 + socioeconomic and behavioral factors (Model 2); and Model 2 + BMI and HbA1c (Model 3, primary). The change in the metformin coefficient from Model 2 to Model 3 was used to estimate the descriptive contribution of metabolic factors [16].

Mortality was modeled using survey-weighted Cox proportional hazards regression, fitted sequentially: crude (Model A); adjusted for demographic, socioeconomic, and metabolic covariates (Model B, primary); Model B plus physical activity and alcohol use (Model C); and Model B with additional adjustment for NLR (Model D) or MLR (Model E). Cardiovascular mortality was analyzed using Models A, B, and D, given the smaller number of events in this subset. Models D and E are exploratory.

NLR was further examined as an independent mortality predictor by categorization into sample-specific tertiles. The metformin–mortality association was then estimated separately within each tertile to explore effect modification.

### Sensitivity Analyses

Five pre-specified sensitivity analyses were performed. First, an active comparator analysis restricted the sample to metformin-monotherapy versus sulfonylurea-monotherapy users (n = 1,056) to reduce confounding by indication. This analysis was applied to cross-sectional outcomes only, given the limited number of events in this subset. Second, multiple imputation was used to address missing data (predictive mean matching, m = 5), and regression estimates were pooled across imputed datasets using Rubin’s rules [17, 18]. However, because the hs-CRP missingness was structural in nature, with an entire cycle absent, the missing-at-random assumption may not hold; imputed hs-CRP results were therefore interpreted with caution. Third, propensity score stratification was conducted using quintiles of a propensity score that included eGFR and concomitant antidiabetic medications [19, 20]. Fourth, overlap weighting was applied to target the average treatment effect in the overlap population [21]. The overlap weights were multiplied by the survey sampling weights and trimmed at the 1st and 99th percentiles to limit the influence of extreme values. Fifth, an exploratory subgroup analysis was conducted across HbA1c categories (<7%, 7–9%, >9%) to examine effect modification. P-values were not adjusted for multiple comparisons across sensitivity and subgroup analyses, and these results are interpreted as hypothesis-generating.

### Ethics

NHANES was approved by the NCHS Research Ethics Review Board, and written informed consent was obtained from all participants. This secondary analysis used de-identified public data and did not require additional institutional review board approval.

### Study Population

The final analytic sample comprised 2,122 adults with self-reported diabetes from NHANES 2013–2018, of whom 1,275 (60.1%) were current metformin users and 847 (39.9%) were non-users (**Table 1**).

**Table 1.** Baseline characteristics of U.S. adults with type 2 diabetes by metformin use, NHANES 2013–2018 (N = 2,122).

| Characteristic | Metformin Users<br>(n=1,275) | Non-Users<br>(n=847) |
| --- | --- | --- |
| **Demographics** |  |  |
| Age, years, mean (SD) | 61.3 (11.7) | 59.9 (15.1) |
| Male, % (SE) | 55.7 (2.3) | 51.5 (2.8) |
| Race/Ethnicity, % (SE) |  |  |
| Mexican American | 10.2 (1.4) | 10.0 (1.9) |
| Other Hispanic | 5.1 (0.6) | 6.5 (0.9) |
| Non-Hispanic White | 61.4 (2.5) | 60.3 (2.6) |
| Non-Hispanic Black | 12.2 (1.5) | 14.2 (1.6) |
| Other/Multi-Racial | 11.2 (1.3) | 8.9 (1.3) |
| **Socioeconomic** |  |  |
| Education, % (SE) |  |  |
| Less than HS | 20.1 (1.8) | 20.4 (2.0) |
| HS diploma/GED | 22.5 (2.1) | 28.6 (2.3) |
| Some college+ | 57.4 (2.1) | 51.0 (2.8) |
| Poverty-Income Ratio, mean (SD) | 2.94 (1.60) | 2.68 (1.66) |
| **Behavioral** |  |  |
| Smoking, % (SE) |  |  |
| Never | 46.8 (2.1) | 51.3 (2.0) |
| Former | 39.1 (2.0) | 34.4 (1.9) |
| Current | 14.0 (1.4) | 14.3 (1.8) |
| Physically active, % (SE) | 38.9 (2.1) | 41.0 (2.2) |
| Alcohol use (any), % (SE) | 71.2 (2.0) | 61.8 (3.0) |
| **Metabolic** |  |  |
| BMI, kg/m<sup>2</sup>, mean (SD) | 32.9 (7.0) | 32.1 (7.1) |
| Obesity (BMI ≥ 30), % (SE) | 62.3 (1.9) | 57.3 (2.8) |
| HbA1c, %, mean (SD) | 7.31 (1.52) | 7.35 (1.91) |
| Diabetes duration, years, mean (SD) | 11.5 (9.7) | 14.9 (12.0) |
| eGFR, mL/min/1.73m<sup>2</sup>, mean (SD) | 87.8 (19.9) | 78.5 (29.4) |
| **Inflammatory Markers** |  |  |
| hs-CRP, mg/L, median (IQR) | 2.30 (1.13, 4.40) | 2.70 (1.34, 5.10) |
| Albumin, g/dL, mean (SD) | 4.17 (0.33) | 4.08 (0.37) |
| NLR, mean (SD) | 2.38 (1.07) | 2.67 (1.74) |
| MLR, mean (SD) | 0.31 (0.13) | 0.33 (0.16) |
| **Concomitant Medications, % (SE)** |  |  |
| Insulin | 17.0 (1.4) | 38.4 (2.0) |
| Sulfonylurea | 28.5 (1.9) | 18.5 (1.9) |
| DPP-4 inhibitor | 12.7 (1.4) | 5.8 (1.0) |
| **Mortality** |  |  |
| All-cause deaths, n | 95 | 124 |
| Cardiovascular deaths, n | 29 | 45 |
| Follow-up, years, median (IQR) | 3.6 (2.4, 5.2) | 3.5 (2.1, 5.2) |

Data are survey-weighted means (SE) for continuous variables and survey-weighted percentages for categorical variables, accounting for the NHANES complex survey design with examination weights pooled across three cycles. Abbreviations: BMI, body mass index; DPP-4, dipeptidyl peptidase-4; eGFR, estimated glomerular filtration rate (2021 CKD-EPI creatinine equation, race-free); HbA1c, glycated hemoglobin; hs-CRP, high-sensitivity C-reactive protein; MLR, monocyte-to-lymphocyte ratio; NLR, neutrophil-to-lymphocyte ratio; PIR, poverty-income ratio.

Survey-weighted estimates indicated that metformin users were slightly older (61.3 vs 59.9 years), more often male (55.7% vs 51.5%), and had higher BMI (32.9 vs 32.1 kg/m²) and eGFR (87.8 vs 78.5 mL/min/1.73 m²). Mean HbA1c was similar between groups (7.31% vs 7.35%). Compared with non-users, metformin users had lower hs-CRP (2.94 vs 3.32 mg/L), higher serum albumin (4.17 vs 4.08 g/dL), and lower NLR (2.38 vs 2.67).

Concomitant insulin use was less frequent among metformin users (17.0% vs 38.4%), whereas sulfonylurea (28.5% vs 18.5%) and DPP-4 inhibitor (12.7% vs 5.8%) co-use was more common. Of the 2,122 participants, 2,116 had valid mortality follow-up data and constituted the prospective mortality cohort; six were excluded because their follow-up time was ≤ 0.

### Metformin Use and Inflammatory Markers

**Table 2** and **Figure 1** summarize the associations between metformin use and inflammatory markers across progressively adjusted models. In the fully adjusted model (Model 3, controlling for age, sex, race/ethnicity, education, PIR, smoking, BMI, and HbA1c), metformin use was significantly associated with higher serum albumin (β = +0.108 g/dL; 95% CI: 0.058, 0.158; p < 0.001), lower NLR (β = −0.352; 95% CI: −0.569, −0.135; p = 0.003), and lower MLR (β = −0.039; 95% CI: −0.056, −0.022; p < 0.001). The association with log(hs-CRP) did not reach statistical significance (β = −0.142; 95% CI: - 0.294, 0.010; p = 0.087), although the direction was consistent with an anti-inflammatory pattern.

**Figure 1.**
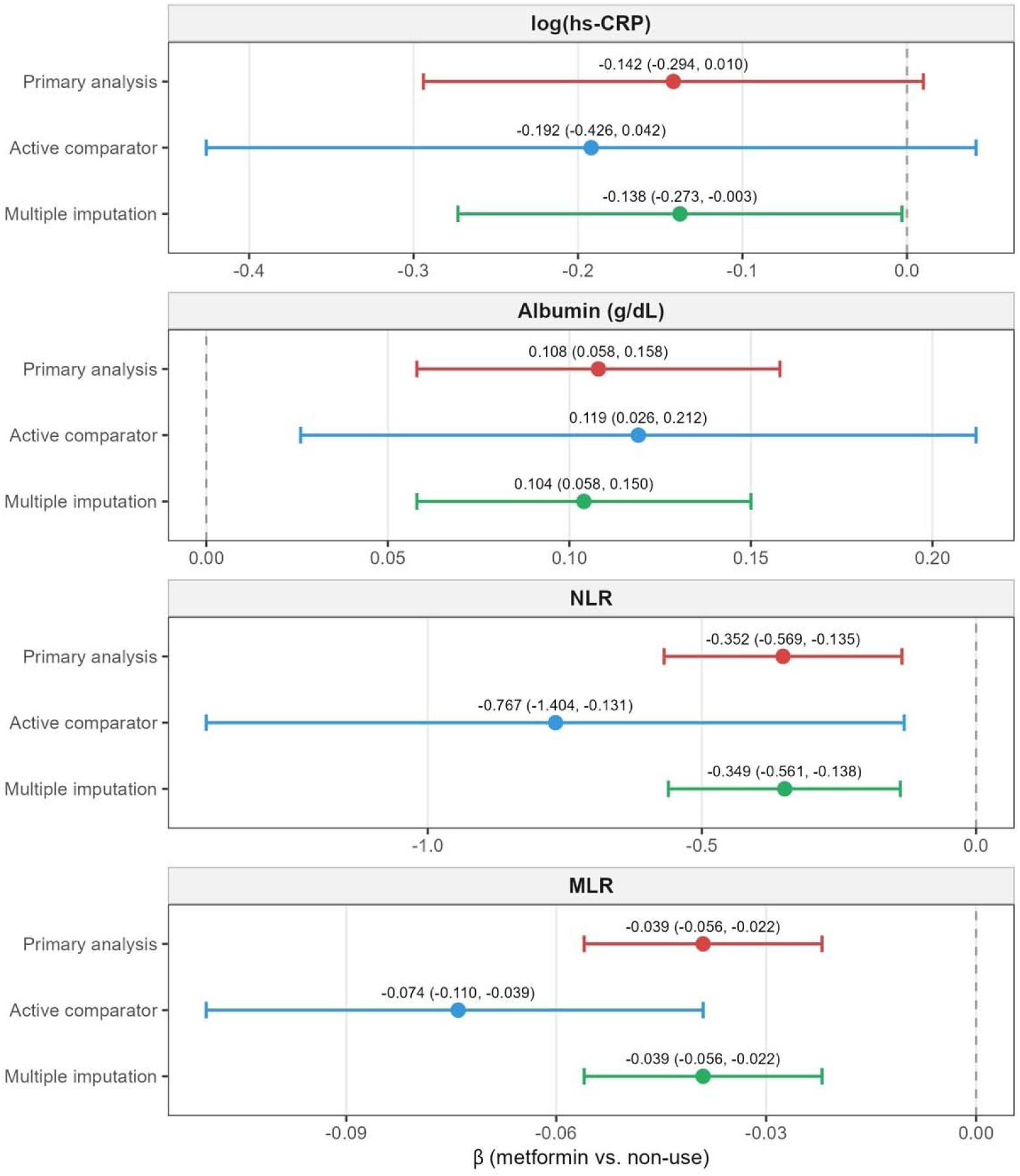
Adjusted associations between metformin use and inflammatory markers across primary, active comparator, and multiple imputation analyses. Forest plot of survey-weighted linear regression coefficients (β) with 95% CIs for log(hs-CRP), serum albumin, NLR, and MLR. Primary analysis: Model 3 (N = 2,122). Active comparator: metformin monotherapy versus sulfonylurea monotherapy (n = 1,056). Multiple imputation: predictive mean matching, m = 5. All models adjusted for age, sex, race/ethnicity, education, PIR, smoking, BMI, and HbA1c. The dashed line indicates the null (β = 0). Abbreviations: hs-CRP, high-sensitivity C-reactive protein; MI, multiple imputation; MLR, monocyte-to-lymphocyte ratio; NLR, neutrophil-to-lymphocyte ratio; PIR, poverty-income ratio.

**Table 2.** Association between metformin use and inflammatory markers, NHANES 2013–2018 (N = 2,122).

| Outcome | Model | Beta | 95% CI | p-value |
| --- | --- | --- | --- | --- |
| log(hs-CRP) | Model 1 | -0.076 | -0.212, 0.060 | 0.261 |
| log(hs-CRP) | Model 2 | -0.09 | -0.247, 0.068 | 0.249 |
| log(hs-CRP) | Model 3 | -0.142 | -0.307, 0.023 | 0.087 |
| Albumin (g/dL) | Model 1 | 0.091 | 0.040, 0.141 | <0.001 |
| Albumin (g/dL) | Model 2 | 0.095 | 0.043, 0.146 | <0.001 |
| Albumin (g/dL) | Model 3 | 0.108 | 0.056, 0.160 | <0.001 |
| NLR | Model 1 | -0.328 | -0.542, -0.113 | 0.004 |
| NLR | Model 2 | -0.344 | -0.569, -0.119 | 0.004 |
| NLR | Model 3 | -0.352 | -0.578, -0.126 | 0.003 |
| MLR | Model 1 | -0.038 | -0.055, -0.021 | <0.001 |
| MLR | Model 2 | -0.039 | -0.056, -0.022 | <0.001 |
| MLR | Model 3 | -0.039 | -0.056, -0.022 | <0.001 |

Survey-weighted linear regression coefficients ( $\beta$ ) with 95% confidence intervals represent the adjusted mean difference in each marker for metformin users versus non-users. Model 1: demographic adjustment (age, sex, race/ethnicity). Model 2: Model 1 plus education, PIR, and smoking. Model 3 (primary): Model 2 plus BMI and HbA1c. The hs-CRP analysis is restricted to participants with available measurements (n = 1,328); hs-CRP was not measured in the 2013–2014 cycle. Abbreviations: CI, confidence interval; hs-CRP, high-sensitivity C-reactive protein; MLR, monocyte-to-lymphocyte ratio; NLR, neutrophil-to-lymphocyte ratio; PIR, poverty-income ratio.

For NLR and MLR, point estimates were similar across all three adjustment levels, with minimal change between Model 2 (pre-BMI/HbA1c adjustment) and Model 3 (post-BMI/HbA1c adjustment). The NLR coefficient shifted only slightly, from −0.344 in Model 2 to −0.352 in Model 3, a change of less than 3%. The metformin-CRP association behaved differently: the inverse estimate strengthened after BMI/HbA1c adjustment, moving from −0.090 in Model 2 to −0.142 in Model 3.

### Change-in-Estimate Analysis

As a descriptive analysis, we compared metformin coefficients from models with and without BMI/HbA1c adjustment for each marker (**Figure 2**).

**Figure 2.**
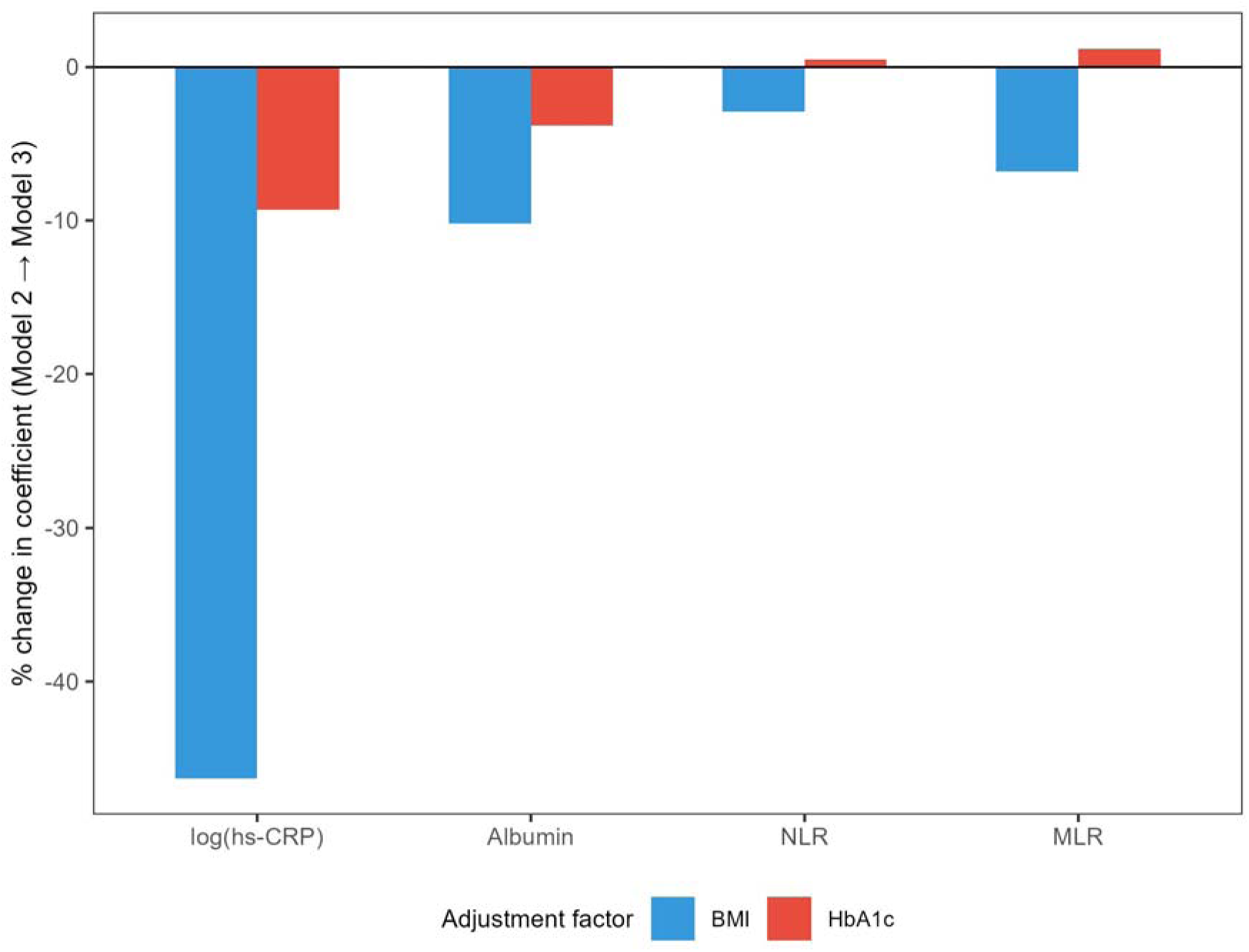
Change in the metformin coefficient after adjustment for BMI and HbA1c (change-in-estimate analysis). Bar chart showing the percent change in the metformin β coefficient from Model 2 (demographic and socioeconomic adjustment) to Model 3 (additionally adjusted for BMI and HbA1c), with separate bars for BMI alone, HbA1c alone, and both combined. Negative values indicate attenuation; for log(hs-CRP), the negative value reflects strengthening of the inverse association after adjustment (i.e., suppression). This analysis is descriptive and does not constitute formal causal mediation. Abbreviations: BMI, body mass index; HbA1c, glycated hemoglobin; hs-CRP, high-sensitivity C-reactive protein; MLR, monocyte-to-lymphocyte ratio; NLR, neutrophil-to-lymphocyte ratio.

For NLR, the percent change attributable to BMI and HbA1c combined was less than 3%; for MLR it was approximately 1%. For albumin, roughly 14% of the association was attenuated after metabolic adjustment. By contrast, the CRP showed that the inverse estimate strengthened by approximately 58% after combined BMI and HbA1c adjustment (from −0.090 to −0.142), with BMI alone accounting for most of this change (46% when added separately).

### Metformin Use and Mortality

During a median follow-up of 3.7 years (range: 0.1–6.9 years), 219 all-cause deaths and 74 cardiovascular deaths were recorded among the 2,116 participants with valid follow-up. Survey-weighted Cox regression results are presented in **Table 3** and Figure 3.

**Figure 3.**
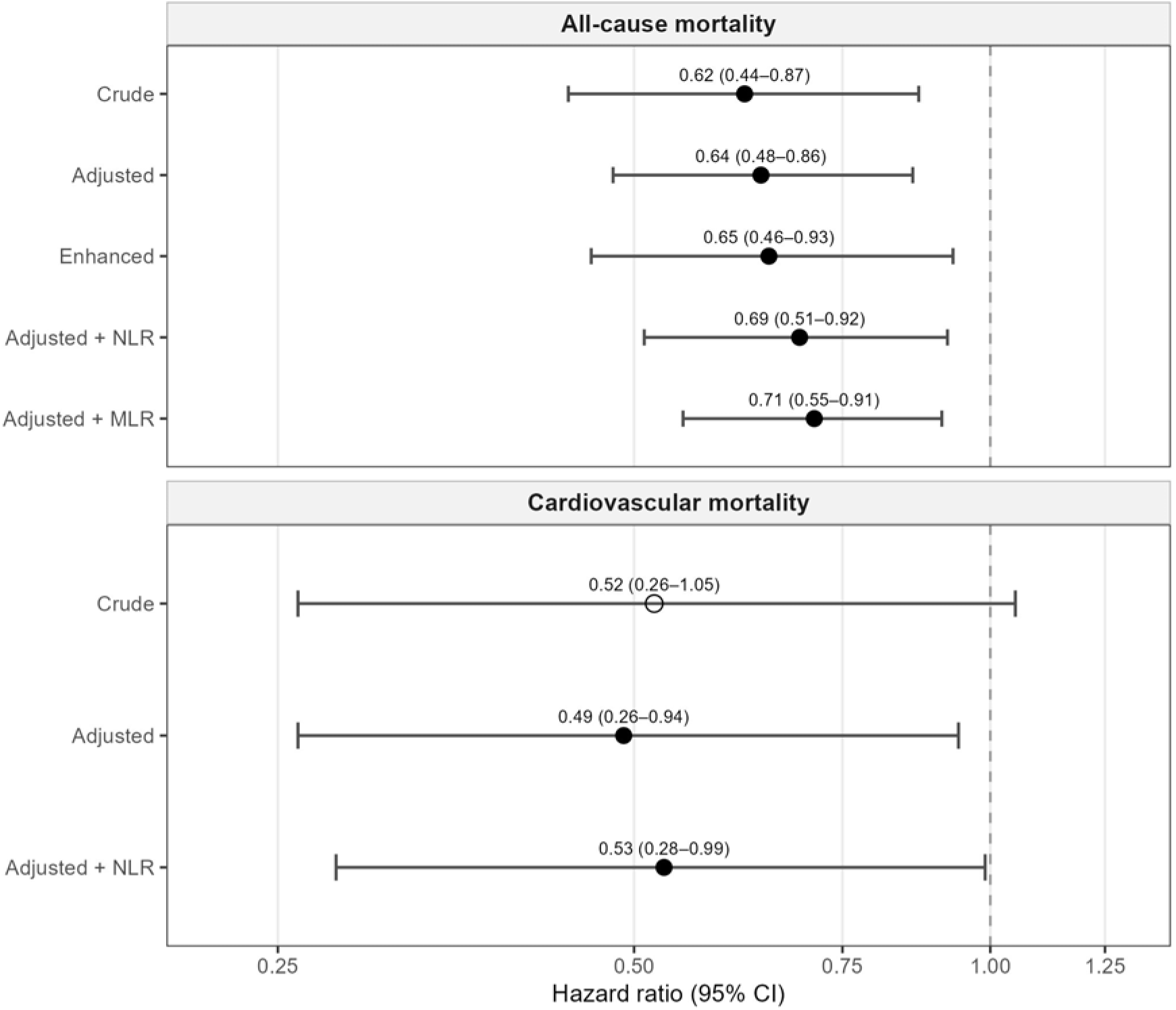
Association between metformin use and mortality across sequential Cox regression models. Forest plot of hazard ratios (95% CIs) for all-cause mortality (upper panel, 219 deaths) and cardiovascular mortality (lower panel, 74 deaths). Models: crude; adjusted (age, sex, race/ethnicity, education, PIR, BMI, HbA1c, smoking); enhanced (adjusted plus physical activity and alcohol); and adjusted plus NLR or MLR. Filled circles denote p < 0.05; open circles denote p ≥ 0.05. The dashed line indicates the null (HR = 1.0). HR axis is plotted on a loge scale with tick labels in the original HR metric. Abbreviations: CI, confidence interval; CV, cardiovascular; HR, hazard ratio; MLR, monocyte-to-lymphocyte ratio; NLR, neutrophil-to-lymphocyte ratio; PIR, poverty-income ratio.

**Table 3.** Association between metformin use and mortality.

| **Model** | **All-cause mortality HR** | **All-cause mortality 95% CI** | **All-cause mortality p** | **Cardiovascular mortality HR** | **Cardiovascular mortality 95% CI** | **Cardiovascular mortality p** |
| --- | --- | --- | --- | --- | --- | --- |
| Model A: Crude | 0.62 | 0.44, 0.87 | 0.006 | 0.52 | 0.26, 1.05 | 0.069 |
| Model B: Adjusted | 0.64 | 0.48, 0.86 | 0.003 | 0.49 | 0.26, 0.94 | 0.031 |
| Model C: Enhanced | 0.65 | 0.46, 0.93 | 0.017 | 0.44 | 0.16, 1.27 | 0.129 |
| Model D: Adjusted + NLR | 0.69 | 0.51, 0.92 | 0.01 | 0.53 | 0.28, 0.99 | 0.047 |
| Model E: Adjusted + MLR | 0.71 | 0.55, 0.91 | 0.006 | 0.54 | 0.29, 1.00 | 0.051 |

Model A: metformin only.
Model B: metformin + age + sex + race/ethnicity + education + PIR + BMI + HbA1c + smoking.
Model C: Model B + physical activity + alcohol use.
Models D/E: Model B + NLR or MLR. As these models condition on potential mediators, results are exploratory and may be subject to collider stratification bias.

For all-cause mortality, metformin use was associated with a significantly lower hazard across all models: crude HR = 0.62 (95% CI: 0.44–0.87; p = 0.006), adjusted HR = 0.64 (95% CI: 0.48–0.86; p = 0.003), and enhanced HR, further adjusting for physical activity and alcohol use, = 0.65 (95% CI: 0.46–0.93; p = 0.017). The association was robust to progressive confounder adjustment, with the hazard ratio remaining stable across models.

For cardiovascular mortality, the crude association did not reach statistical significance (HR = 0.52; 95% CI: 0.26–1.05; p = 0.069), whereas the adjusted model yielded HR = 0.49 (95% CI: 0.26–0.94; p = 0.031). The enhanced model, Model C, yielded HR = 0.44 (95% CI: 0.16–1.27; p = 0.129). However, the widened confidence interval reflects limited statistical power(74 events and 12 parameters), and this estimate should be interpreted with caution. Covariate adjustment improved precision without substantially altering the point estimate, from 0.52 in the crude model to 0.49 in the adjusted model, suggesting the robustness.

When NLR was added to the adjusted all-cause mortality model, the metformin HR shifted from 0.64 to 0.69 (95% CI: 0.51–0.92; p = 0.010), representing a modest attenuation. A similar pattern was observed with MLR, with the HR shifting from 0.64 to 0.71 (95% CI: 0.55–0.91; p = 0.006). For cardiovascular mortality, adding NLR shifted the HR from 0.49 to 0.53 (95% CI: 0.28–0.99; p = 0.047), and adding MLR yielded HR = 0.54 (95% CI: 0.29–1.00; p = 0.051).

### NLR and MLR as Independent Mortality Predictors

In a survey-weighted Cox model adjusting for age, sex, race/ethnicity, education, PIR, BMI, HbA1c, smoking, and metformin use, NLR analysed in sample-specific tertiles was independently associated with all-cause mortality (**Figure 4**). Compared with the lowest tertile (T1), the highest tertile (T3) carried a significantly elevated hazard (HR = 1.95; 95% CI: 1.41–2.68; p < 0.001), whereas the middle tertile showed a non-significant elevation (HR = 1.11; 95% CI: 0.74–1.67; p = 0.620). MLR was likewise an independent predictor (HR = 1.25 per 0.1-unit increase; 95% CI: 1.17–1.35; p < 0.001).

**Figure 4.**
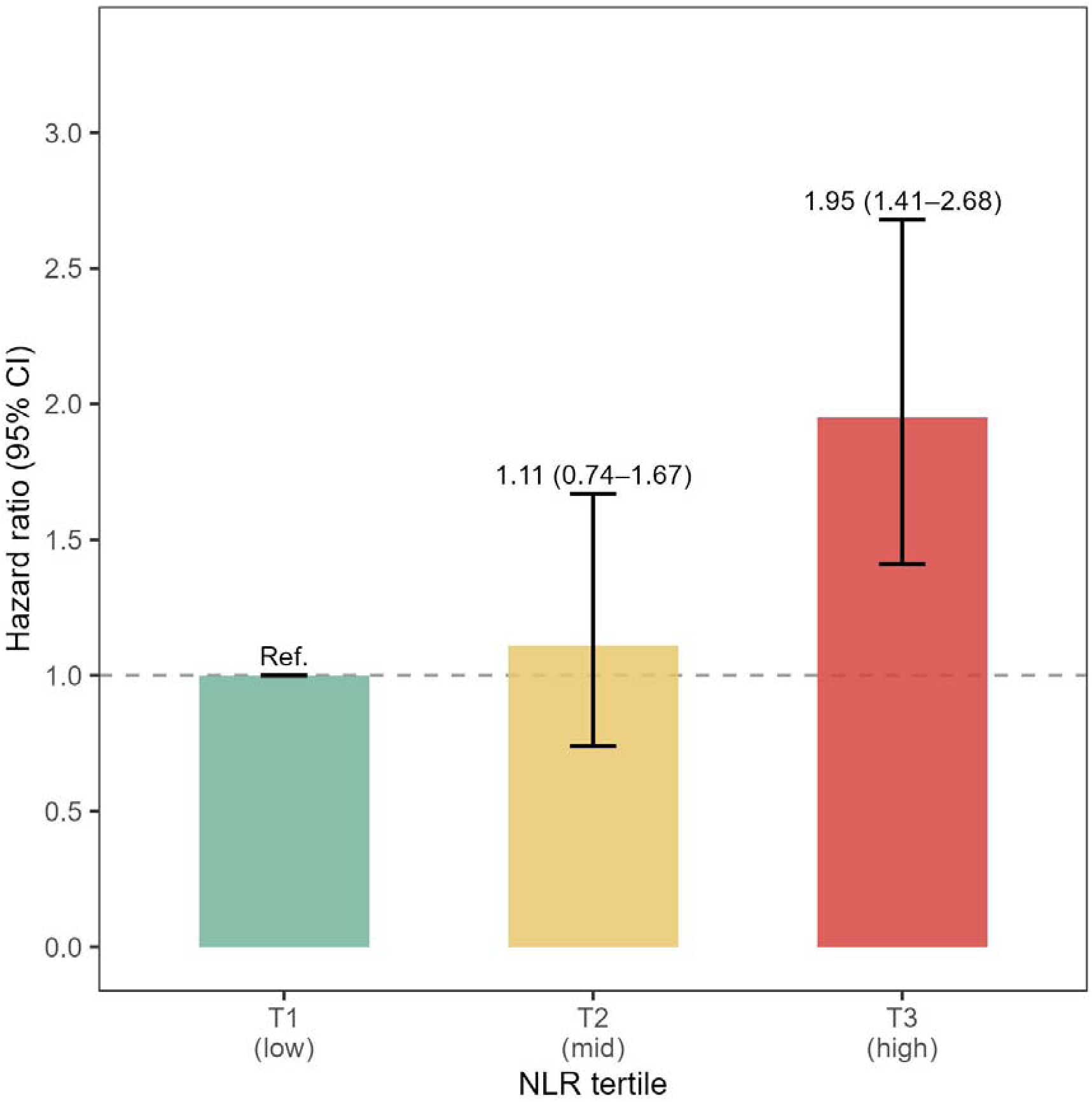
All-cause mortality by NLR tertile in adults with type 2 diabetes. Survey-weighted Cox regression hazard ratios (95% CIs) for all-cause mortality by NLR tertile (T1 = reference; cut-points 1.74 and 2.55; n = 683, 677, and 678 in T1, T2, T3). Adjusted for age, sex, race/ethnicity, education, PIR, BMI, HbA1c, smoking, and metformin use. The dashed line indicates the null (HR = 1.0). Abbreviations: CI, confidence interval; HR, hazard ratio; NLR, neutrophil-to-lymphocyte ratio; PIR, poverty-income ratio.

A formal interaction test (metformin × NLR, continuous) was non-significant (p = 0.905), providing no evidence of effect modification by baseline NLR level.

### Sensitivity Analyses

#### Active Comparator

In the active comparator analysis restricted to metformin monotherapy versus sulfonylurea monotherapy (n = 1,056), metformin was associated with lower NLR (β = - 0.767; 95% CI: −1.404, −0.131; p = 0.025), lower MLR (β = −0.074; 95% CI: −0.110, - 0.039; p < 0.001), and higher albumin (β = +0.119; 95% CI: 0.026, 0.212; p = 0.018) (**Figure 1)**. The NLR point estimate was approximately twice as large as in the primary analysis, possibly reflecting reduced confounding by indication in this head-to-head comparison.

#### Multiple Imputation

After multiple imputation (m = 5, predictive mean matching), results for albumin (β = +0.104; p < 0.001), NLR (β = −0.349; p = 0.001), and MLR (β = −0.039; p < 0.001) were essentially unchanged from the complete-case analysis. The log(hs-CRP) association, which did not reach significance in the primary analysis (p = 0.087), became nominally significant after imputation (β = −0.138; 95% CI: −0.273, −0.003; p = 0.046). However, the high fraction of missing information (FMI = 0.40) indicates that this estimate is substantially driven by the imputation model.

#### Propensity Score Methods

Propensity score stratification (quintiles) yielded directionally consistent results: albumin (pooled β = +0.076; p = 0.003) and log(hs-CRP) (pooled β = −0.142; p = 0.046) remained significant, whereas NLR was attenuated (pooled β = −0.274; p = 0.058). Overlap weighting similarly confirmed albumin (+0.084; p = 0.003) and NLR (−0.270; p = 0.043), with log(hs-CRP) remaining borderline (−0.129; p = 0.100). The variability in NLR significance across methods (PS stratification p = 0.058; overlap weighting p = 0.043) suggests that the effect size lies near the detection threshold in this sample, although the directional consistency supports the primary findings.

#### HbA1c Subgroup Analysis

In an exploratory subgroup analysis stratified by HbA1c (**Figure S2**), the metformin–inflammation associations were most consistently observed in the 7–9% subgroup, where all four markers showed significant associations (log[hs-CRP]: β = −0.310, p = 0.030; albumin: +0.176, p < 0.001; NLR: −0.614, p < 0.001; MLR: −0.047, p < 0.001). In the < 7% and > 9% subgroups, only albumin and MLR remained significant.

#### Enhanced Confounder Adjustment

Additional adjustment for physical activity, alcohol use, and diabetes duration **(Table 2)** did not materially alter the cross-sectional results: albumin remained significant (β = +0.084; p = 0.028), as did NLR (β = −0.346; p = 0.027) and MLR (β = −0.036; p = 0.003).

## Discussion

In this nationally representative sample of U.S. adults with diabetes, metformin use was associated with a more favorable cellular immune-inflammation profile, lower NLR and MLR, and higher serum albumin, independent of demographic, socioeconomic, and metabolic factors. These associations persisted across progressively adjusted models, in an active comparator comparison against sulfonylurea monotherapy, and across multiple sensitivity analyses. The hs-CRP association was directionally consistent but less robust. Metformin use was also associated with lower all-cause and cardiovascular mortality, and inclusion of NLR or MLR in the mortality model produced a modest attenuation of this association. Elevated NLR was itself independently associated with mortality, following a threshold rather than linear dose–response pattern.

Our NLR findings are consistent with a Scottish clinical cohort comparing metformin with sulfonylurea users [11], extending that observation to a diverse, population-based U.S. sample with prospective mortality follow-up. The independent NLR–mortality relationship is consistent with prior NHANES-based evidence, to which our analysis adds the medication dimension by incorporating metformin use and exploring potential heterogeneity across NLR strata [6, 22]. The albumin finding aligns with recent evidence identifying albumin as relevant to metformin-associated mortality differences in elderly hypertensive patients; we extend this to the broader diabetic population and situate it alongside CBC-derived cellular indices not previously examined in this context. Population-level evidence for MLR remains sparse, and our findings contribute to its emerging characterization as a metformin-responsive index of monocyte-driven inflammation [11, 12].

The differential attenuation pattern across markers is informative. The metformin–NLR and metformin–MLR associations were essentially unaffected by adjustment for adiposity and glycemic control, suggesting that they do not simply reflect metabolic differences between users and non-users. In contrast, the hs-CRP association was largely attenuated by BMI adjustment, indicating that CRP differences between groups predominantly reflect adiposity-related processes rather than a distinct immune–inflammatory pathway. Albumin showed an intermediate pattern, consistent with its dual role as both an inflammatory and nutritional marker.

This divergence is biologically plausible. Experimental studies have shown that metformin directly modulates neutrophil and monocyte function via AMPK activation and mitochondrial pathways [3, 4, 23], which plausibly correspond to cellular immune indices rather than to hepatic acute-phase responses more tightly coupled to adiposity signalling [10]. The consistency of this pattern across primary, active comparator, and propensity score analyses strengthens the inference that metformin’s associations with cellular immune indices are unlikely to be fully explained by metabolic differences.

The modest attenuation of metformin’s mortality association upon inclusion of NLR or MLR is compatible with cellular inflammation reflecting one component of the overall mortality association, rather than a dominant mediating pathway. This interpretation is necessarily descriptive: with exposure, markers, and outcomes captured cross-sectionally for the marker analysis and prospectively for mortality, temporal ordering cannot be resolved and formal causal mediation is not estimable. Although a formal test for effect modification was non-significant, descriptive HR patterns across NLR strata may warrant exploration in larger prospective studies with sufficient power for interaction testing.

Strengths include the nationally representative design, the combined cross-sectional and prospective framework, and four complementary sensitivity approaches (active comparator, multiple imputation, propensity score stratification, overlap weighting) addressing distinct sources of bias with broadly consistent results. The markers examined are derived from routine clinical panels, enhancing translational relevance.

Several limitations warrant note. Residual confounding is possible from unmeasured factors such as diet, infection burden, or concomitant medications. We used a prevalent-user rather than new-user design, as longitudinal prescription data are unavailable in NHANES; this approach is susceptible to depletion-of-susceptibles and survivor effects that may bias associations toward apparent benefit [24], and metformin dose and duration could not be assessed. The relatively short follow-up limited power for cardiovascular mortality and may be insufficient for inflammation-mediated pathways, whose consequences accrue over longer horizons. Self-reported diabetes and medication use introduce potential misclassification, and hs-CRP had substantial missingness due to a mid-cycle protocol change.

The observation that metformin’s associations with CBC-derived cellular indices persist after adjustment for adiposity and glycaemic control supports further investigation of non-glycaemic pathways as contributors to metformin’s long-recognised clinical benefits. The differential pattern across markers suggests that future studies may benefit from distinguishing between cellular and acute-phase inflammatory markers rather than treating “inflammation” as a unitary construct. Prospective studies with repeated measurements, and where feasible interventional designs, are needed to establish temporal ordering and causal relevance. As routinely available and inexpensive indices, NLR and MLR represent pragmatic candidates for inclusion in such future investigations.

## Data Availability

NHANES was approved by the NCHS Research Ethics Review Board, and written informed consent was obtained from all participants. This secondary analysis used de-identified public-use data and did not require additional institutional review board approval.

https://wwwn.cdc.gov/nchs/nhanes/.

https://www.cdc.gov/nchs/data-linkage/mortality-public.htm.

## Data Availability

All data used in this study are publicly available. NHANES survey data can be accessed at https://wwwn.cdc.gov/nchs/nhanes/. The NHANES Public-Use Linked Mortality Files are available at https://www.cdc.gov/nchs/data-linkage/mortality-public.htm. Analytic code is available from the corresponding author upon reasonable request.

## Author Contributions

YC and HS conceptualised the study. YC conducted the data analysis and drafted the manuscript. JG, YW, and YX contributed to data interpretation and critical revision of the manuscript. HS supervised the study and provided critical revision. All authors approved the final version of the manuscript.

## Supplementary material

**Supplementary table 1.**
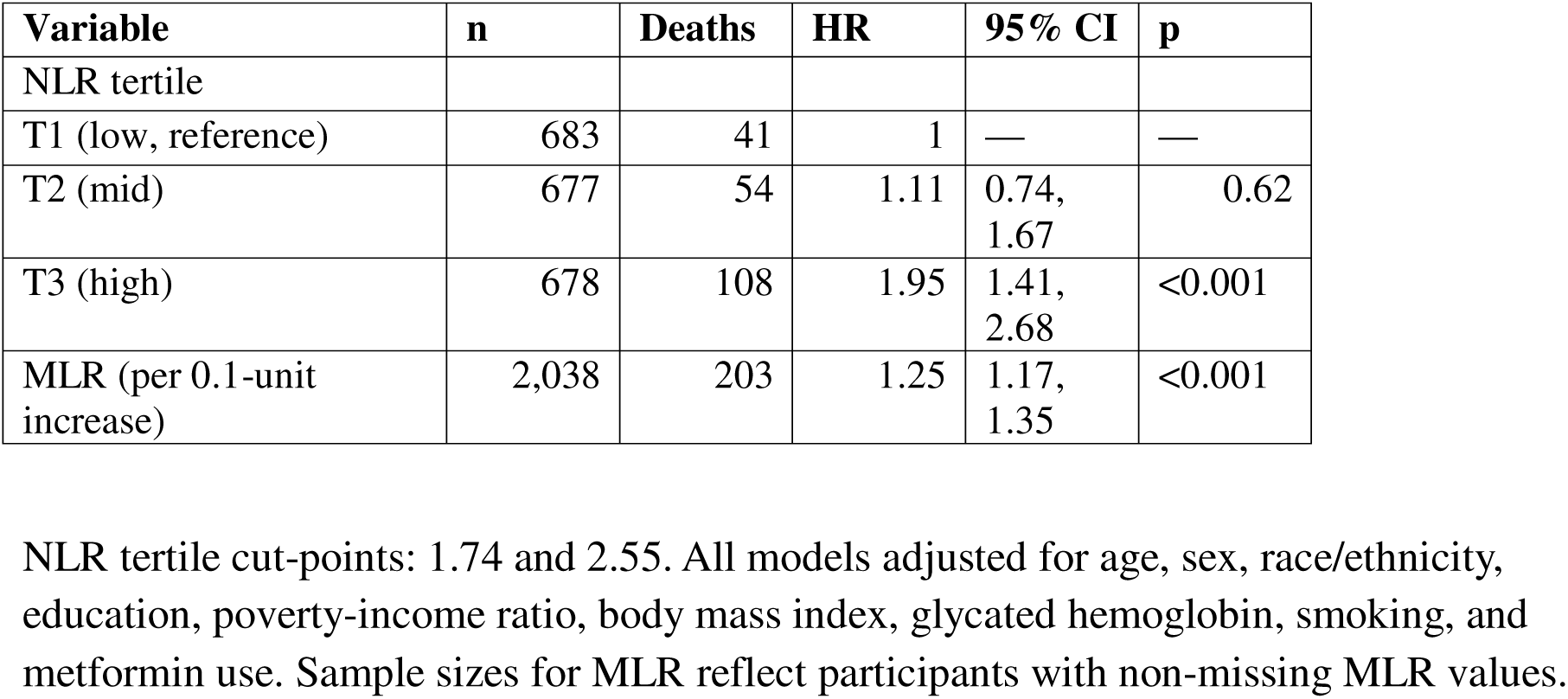
NLR and MLR as Independent Predictors of All-Cause Mortality.

| Variable | n | Deaths | HR | 95% CI | p |
| --- | --- | --- | --- | --- | --- |
| NLR tertile |  |  |  |  |  |
| T1 (low, reference) | 683 | 41 | 1 | — | — |
| T2 (mid) | 677 | 54 | 1.11 | 0.74,<br>1.67 | 0.62 |
| T3 (high) | 678 | 108 | 1.95 | 1.41,<br>2.68 | <0.001 |
| MLR (per 0.1-unit increase) | 2,038 | 203 | 1.25 | 1.17,<br>1.35 | <0.001 |

NLR tertile cut-points: 1.74 and 2.55. All models adjusted for age, sex, race/ethnicity, education, poverty-income ratio, body mass index, glycated hemoglobin, smoking, and metformin use. Sample sizes for MLR reflect participants with non-missing MLR values.

**Figure S1.**
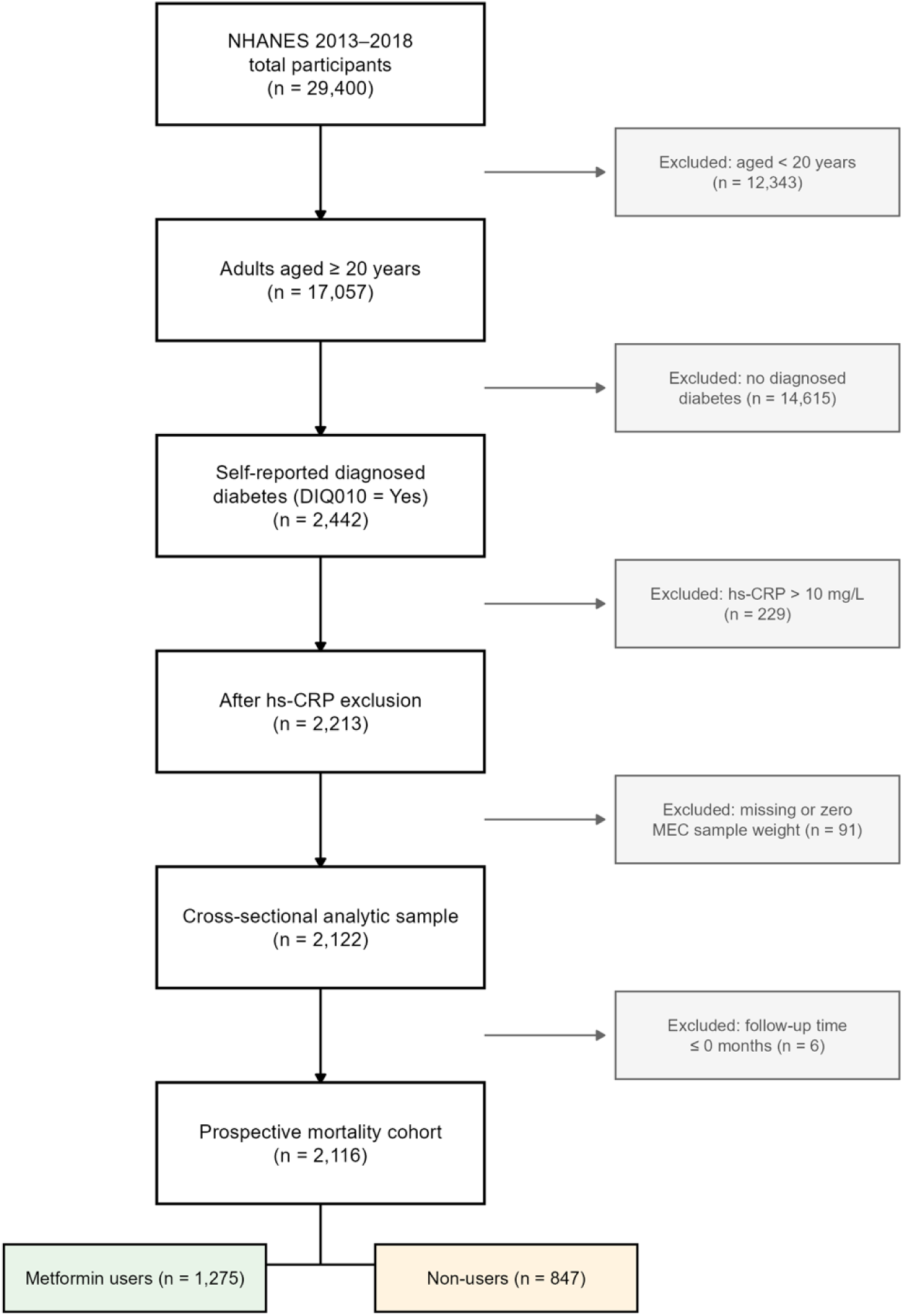
Inclusion and exclusion of participants.

**Figure S2.**
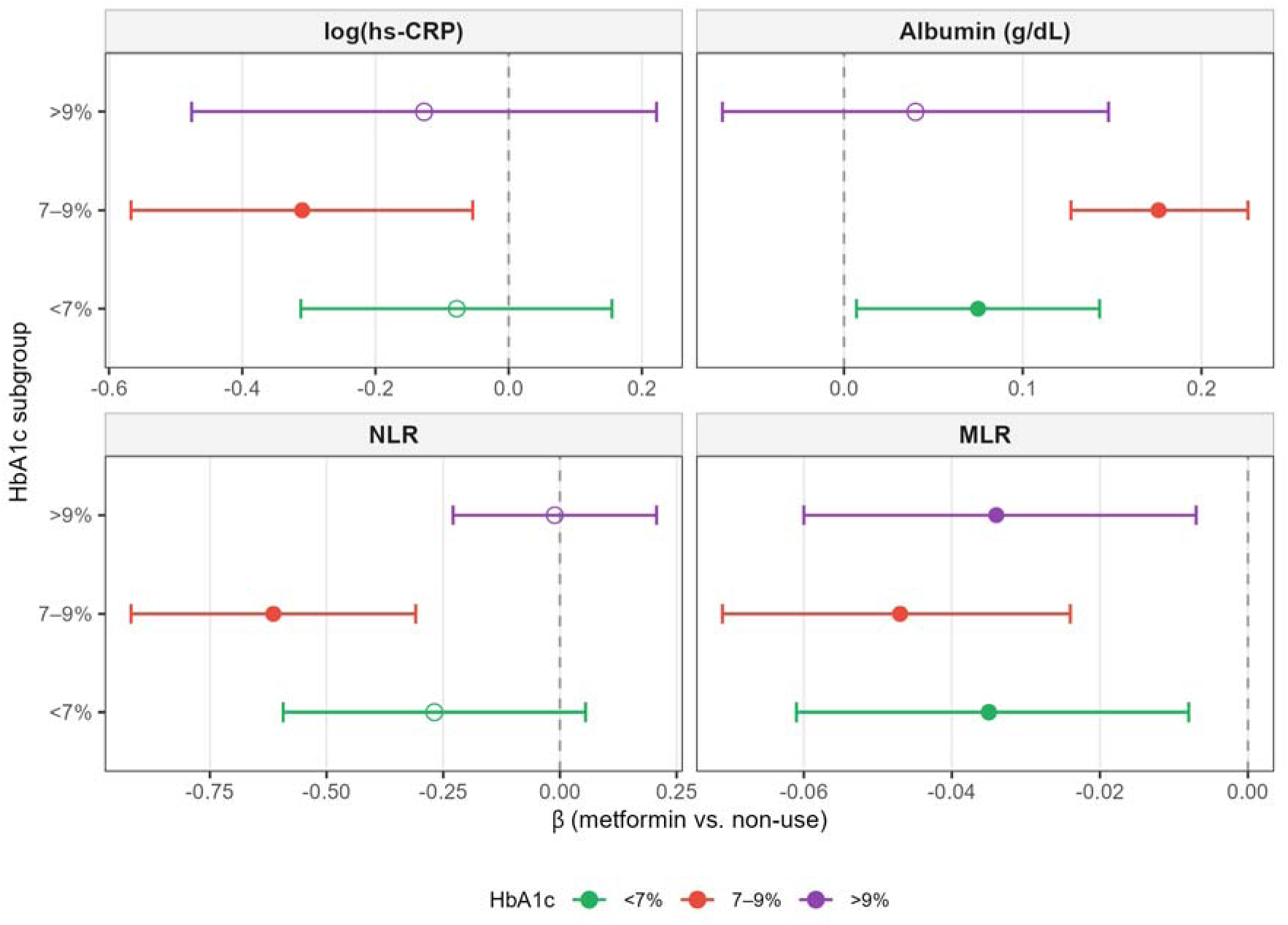
Association between metformin use and inflammatory markers, stratified by HbA1c subgroup. Forest plot of survey-weighted linear regression coefficients (β) with 95% CIs for log(hs-CRP), serum albumin, NLR, and MLR within HbA1c subgroups (<7%, 7–9%, >9%). Models adjusted for age, sex, race/ethnicity, education, PIR, smoking, and BMI; HbA1c was omitted from the covariate set as it defines the strata. Filled circles denote p < 0.05; open circles denote p ≥ 0.05. The dashed line indicates the null (β = 0). This analysis was pre-specified as exploratory; p-values were not adjusted for multiple comparisons. Abbreviations: HbA1c, glycated hemoglobin; hs-CRP, high-sensitivity C-reactive protein; MLR, monocyte-to-lymphocyte ratio; NLR, neutrophil-to-lymphocyte ratio; PIR, poverty-income ratio.

